# Factors Associated With Regional Differences in Healthcare Quality for Patients With Acute Myocardial Infarction in Japan

**DOI:** 10.1101/2022.05.20.22275402

**Authors:** Shusuke Watanabe, Jung-ho Shin, Etsu Goto, Susumu Kunisawa, Yuichi Imanaka

## Abstract

**Objectives:** Regional medical coordination is essential for health-system reform in Japan, and the quality of healthcare at the regional level is of great interest. Regional differences in the quality of healthcare for cardiovascular diseases have been pointed out in previous research, and we aimed to clarify regional factors that influence the quality.

**Design:** Insurance claims database study.

**Setting:** Patients of acute care in Japan.

**Participants:** Japanese patients included in the national Diagnosis Procedure Combination (DPC) databases who admitted to hospitals with diagnosis of acute myocardial infarction (AMI) from April 2016 to March 2019.

**Main outcome and measures:** Using the national DPC databases, risk-adjusted mortality (RAM) in each secondary medical area (SMA) was derived as an indicator for the quality of the healthcare for patients with AMI. Factors associated with RAM were analysed using the partial least squares (PLS) regression model.

**Results:** There is a wide variation in RAM in the SMAs; the maximum value was 0.593 and the minimum value was 1.445. The PLS regression identified two components positively correlated with RAM. The first component (Component 1) positively correlates with the proportion of the elderly within the population and negatively correlates with the number of medical facilities per area, the population density, and the intra-SMA access to centres with a high volume of emergency percutaneous coronary interventions (ePCI). The second component (Component 2) positively correlates with the number of medical facilities per area and negatively correlates with the number of physicians per person and the intra-SMA access to centres with a high volume of ePCI.

**Conclusion:** There was wide regional variation in the quality of in-hospital AMI treatments. Results suggested the degree of rurality, the sufficiency of medical resources, the access to high-volume ePCI centres, and coordination of healthcare delivery were associated with healthcare quality for AMI patients.

## INTRODUCTION

Japanese healthcare is essentially financed by social health insurance (SHI) but is mainly provided by private bodies.[1] To improve coordination between medical facilities owned by various bodies, prefectural governments are responsible for establishing regional medical plans.[2] Such plans set goals and action plans to improve regional health systems, and each medical facility should make efforts to cooperate with said plans.[2]

Reforms to promote regional medical coordination have become more important in recent years[3] due to the rapid changes in population structure.[4] An aging population presents changes in medical needs and a decrease in the working population, resulting in the constraint of medical resources.[5, 6] Consequently, reform intends to accelerate regional coordination to improve efficiency.

Along with improving efficiency, quality assurance is a key goal of the regional medical plans.[2] Healthcare quality is defined as “the degree to which health services for individuals and populations increase the likelihood of desired health outcomes and are consistent with current professional knowledge,”[7] and this definition can be applied to a regional level. In terms of the assessment of healthcare quality, among many other indicators, risk-adjusted mortality (RAM) is a widely-used indicator directly related to patient outcomes.[8, 9]

While regional differences in healthcare quality for acute diseases are of great interest,[10–14] little is known about the factors associated with the quality at a regional level. Regarding acute cardiovascular diseases, the presence of more cardiologists[15] and a lower time from the onset of disease to arrival at the hospital[16] were suggested to have a positive association with outcomes at the hospital level. However, at the regional level, more cardiologists may not positively impact the quality, unless cardiologists can take advantage of medical facilities for acute care, and unless patients requiring specialised care have smooth access to hospitals of sufficient quality. The outcomes of cardiovascular diseases have also been suggested to have regional differences, associated with social and healthcare factors.[17, 18] Behavioural risks and medical spending are reported to have a positive impact on outcomes,[17, 18] though as these studies directly focused on all of the residents’ outcomes, it is uncertain whether the suggested factors are associated with healthcare quality or the residents’ health conditions before arriving at the hospital.

As mentioned in the previous sections, many medical and social factors, including the residents’ features, are associated with the quality of healthcare for cardiovascular diseases. However, the relationship between the regional factors and the structure to influence the quality are ambiguous. These many regional factors are intercorrelated. This study aims to clarify the regional differences in healthcare quality for acute myocardial infarctions (AMIs) and to disentangle the regional factors associated with healthcare quality.

## METHODS

### Secondary Medical Areas in Japan

In the regional medical plans, secondary medical areas (SMAs) are designated units that provide general in-hospital healthcare.[2, 19] As of January 2022, there are 335 secondary medical areas. In Japan, the 52 area units for the management of more advanced treatments are designated as tertiary medical areas, while more primary treatments are managed within smaller areas, across approximately 1,700 districts.[19] We selected secondary medical areas as units of analysis given they are the basic units in the coordination of in-hospital care. For consistency of analysis, we utilised the area setting as of January 2022.

### Data Sources

We utilised data from three databases, adding to publicly open data. The first two databases consisted of Diagnosis Procedure Combination (DPC) data, one of which is managed by the Ministry of Health, Labour, and Welfare (MHLW), hereinafter referred to as the MHLW DPC database. DPC data are used for the Diagnosis Procedure Combination/Per-Diem Payment System (DPC/PDPS)—a reimbursement system set up for acute care—and are also used to improve systems and policies in Japan.[20] Hence, the data include patient clinical information such as disease severity, adding to the information of clinical procedures.[20] From this MHLW DPC database, aggregated data can be obtained for research purposes.

The second database utilised was that of the nationwide database of DPC data submitted from participating hospitals for research, hereinafter referred to as the nationwide DPC research database. While not all hospitals included in the MHLW DPC database participated in the nationwide DPC research database, the individual data are available for research.

The third database is the National Database of Health Insurance Claims and Specific Health Check-ups of Japan (NDB), which is a national database of SHI claims data and includes information on all the medical procedures covered by the SHI in Japan.[21, 22]

### Regional Factors Associated with the Quality of AMI Healthcare

The factors associated with differences in the indicator values of healthcare quality for AMI patients across regions were analysed using the partial least squares (PLS) regression model with regional variables.

#### Regional Quality Indicators of Healthcare for AMI Patients

As a quality indicator of care for patients with AMIs, RAM was derived as below. First, based on the data from the nationwide DPC research database, a prediction model for in-hospital mortality of patients admitted with diagnoses of AMI was produced. The employed predictors at admission were sex, age, body mass index (BMI), Killip classification, and the Japan Coma Scale. Data of patients admitted between July 2010 and March 2018 with diagnoses of AMIs were included for the establishment of the model. A multilevel logistic regression model with random intercepts of hospital identifiers was employed to account for clustering within the facilities.[23] Second, aggregated data of case volume, expected deaths, and observed deaths of in-hospital AMI patients in each SMA from fiscal year (FY) 2016 to FY 2018 (from April 2016 to March 2019) were obtained from the MHLW DPC database. The expected deaths were calculated by the derived prediction model. Third, the ratio of expected deaths to observed deaths in each area from FY 2016 to FY 2018 was calculated as RAM, a regional quality indicator of the AMI treatment, with risk adjustment through indirect standardization.

#### Regional Variables

The variables of regional medical resources, resident features, and basic geographical features are included as explanatory variables. The details of employed variables are presented in Supplementary Table 1. All the variables were gathered from publicly available data, excluding the indicator of intra-SMA access to centres with a high volume of emergency percutaneous coronary interventions (ePCI), hereinafter, referred to as the Indicator of Access. This indicator was derived using data extracted from the NDB, which includes all the medical procedures covered by the SHI. The median case number of emergency PCIs of hospitals where at least one ePCI occurred from FY 2016 to FY 2018 was set as the threshold. Then, the hospitals with case numbers over the threshold were defined as the high-volume ePCI centres, and an ePCI share of high-volume ePCI centres was calculated as the indicator.

#### Imputation of Missing Death Numbers in SMAs

For privacy reasons, when the observed deaths were smaller or equal to nine, the exact numbers were masked in the available data, resulting in missing RAM values. To deal with these non-random missing values in dependent variables, imputation was performed as follows. First, the association between AMI case volumes and crude mortality was analysed. Then, based on the association, the crude mortality for SMAs with missing values was estimated and imputed. Specifically, the weighted mean of the observed crude mortality in SMAs was used for the estimation, where case volumes were under the 90^th^ percentile of the distribution of case volume of SMAs with missing values. Missing observed deaths were calculated using the imputed crude mortality, and when the estimated number of deaths was larger than the possible maximum value of nine, nine was imputed. Finally, RAM was derived from the imputed observed deaths.

#### Regression Models

To analyse the association between regional factors and the quality of healthcare for AMI patients, regression analyses with the dependent variable of RAM were performed. Since the regional factors of interest in this study were intended to be interdependent, the PLS regression was undertaken as the main analysis. PLS regression was employed for epidemiological data with multicollinearity.[24–28] Ordinary least squares (OLS) regression was performed to examine multicollinearity.

#### Statistical Analyses

The number of components in the PLS regression was set by evaluating the cumulative explained variance of the dependent variable by components. To avoid explanatory variables with large absolute values dominating the analysis, data centring and scaling were performed before PLS regression.[24]

SAS software version 9.4 (SAS Institute Inc., Cary, NC) was used for setting up the prediction model for AMI patient mortality. PLS regression, other statistical analyses, and plotting were carried out using R version 4.0.5 (R Foundation for Statistical Computing, Vienna, Austria) with the R package “*pls*” version 2.8-0.[29]

#### Sensitivity Analyses

In the PLS regression, sensitivity analyses were undertaken for the imputation of missing values of observed deaths and the Indicator of Access. For the imputation of missing values of observed deaths, in the first sensitivity analysis, the crude mortality used for estimation was the weighted mean of the mortality in SMAs, where case volumes were under the maximum value, rather than the 90^th^ percentile, of the case volumes of SMAs with missing values. The second sensitivity analysis was undertaken, limited to the SMAs where exact observed deaths were obtained.

For the Indicator of Access, in the first sensitivity analysis, SMAs where no ePCIs were performed were removed. In these areas, access was not considered to be high but the coordination between hospitals was hardly considered as completed in the SMAs. In the second sensitivity analysis, the indicator was set to one if there was only one hospital that undertook ePCIs in a SMA, even when case volumes of those hospitals were under the threshold. These hospitals are considered to have a central role in the SMAs.

## RESULTS

### Quality Indicators of AMI Treatment and the Characteristics of Medical Areas

For the establishment of the prediction model, 394,087 patients were included and there were 55,727 in-hospital deaths. The c-statistic of the derived prediction model was 0.9177. The observed and predicted deaths for each SMA were made public on the website of the Department of Healthcare Economics and Quality Management, Graduate School of Medicine, Kyoto University (http://med-econ.umin.ac.jp/fH9VdGtT/).

Table 1 shows the characteristics of the SMAs. The median area and population of the SMAs were 855.66 km^2^ and 224,369, respectively. There were missing RAM values for 103 SMAs and in 32 SMAs there were no facilities undertaking ePCIs.

**Table 1.**
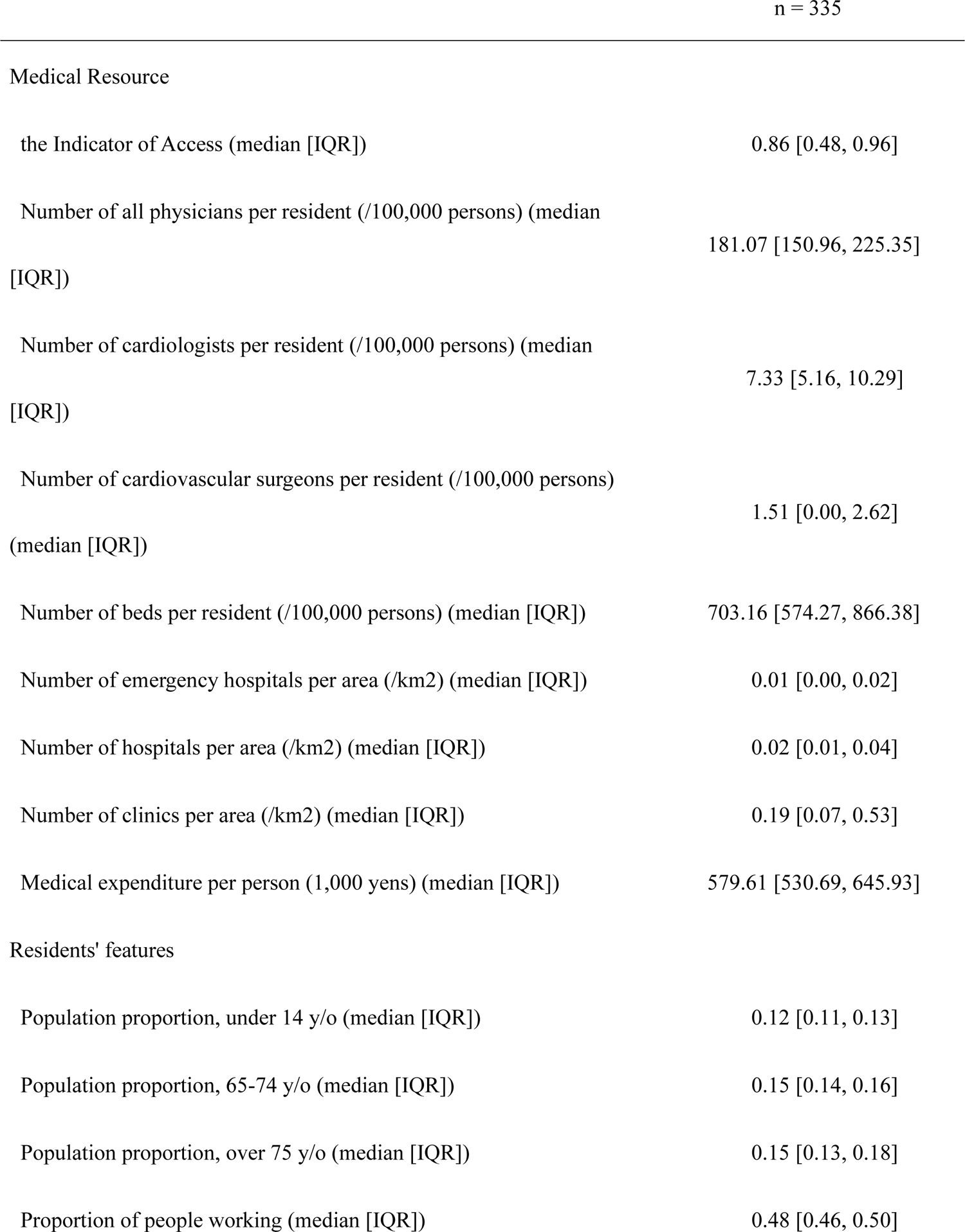
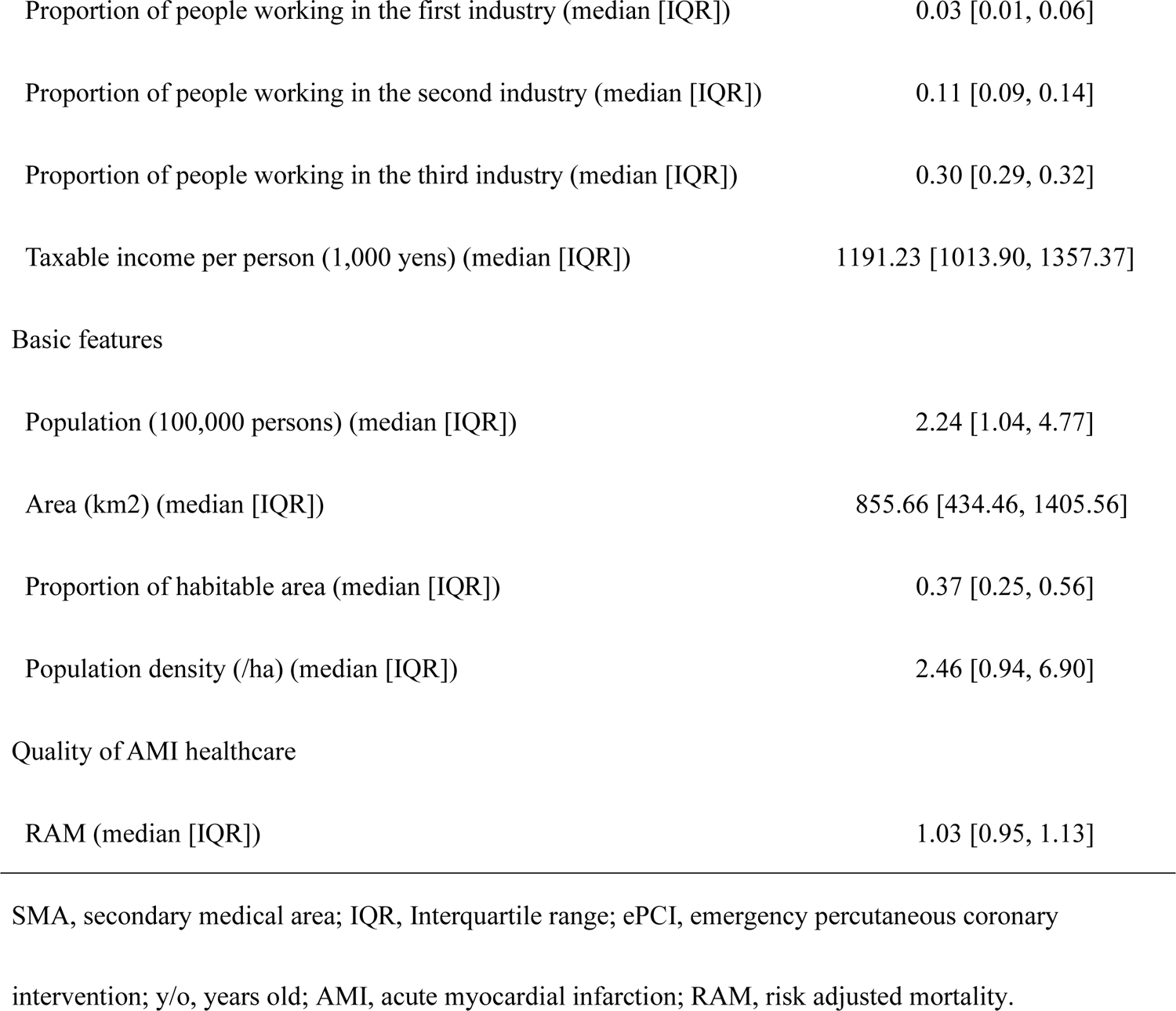
Characteristics of SMAs

|  | n = 335 |
| --- | --- |
| Medical Resource |  |
| the Indicator of Access (median [IQR]) | 0.86 [0.48, 0.96] |
| Number of all physicians per resident (/100,000 persons) (median [IQR]) | 181.07 [150.96, 225.35] |
| Number of cardiologists per resident (/100,000 persons) (median [IQR]) | 7.33 [5.16, 10.29] |
| Number of cardiovascular surgeons per resident (/100,000 persons) (median [IQR]) | 1.51 [0.00, 2.62] |
| Number of beds per resident (/100,000 persons) (median [IQR]) | 703.16 [574.27, 866.38] |
| Number of emergency hospitals per area (/km<sup>2</sup>) (median [IQR]) | 0.01 [0.00, 0.02] |
| Number of hospitals per area (/km<sup>2</sup>) (median [IQR]) | 0.02 [0.01, 0.04] |
| Number of clinics per area (/km<sup>2</sup>) (median [IQR]) | 0.19 [0.07, 0.53] |
| Medical expenditure per person (1,000 yens) (median [IQR]) | 579.61 [530.69, 645.93] |
| Residents' features |  |
| Population proportion, under 14 y/o (median [IQR]) | 0.12 [0.11, 0.13] |
| Population proportion, 65-74 y/o (median [IQR]) | 0.15 [0.14, 0.16] |
| Population proportion, over 75 y/o (median [IQR]) | 0.15 [0.13, 0.18] |
| Proportion of people working (median [IQR]) | 0.48 [0.46, 0.50] |
| Proportion of people working in the first industry (median [IQR]) | 0.03 [0.01, 0.06] |
| Proportion of people working in the second industry (median [IQR]) | 0.11 [0.09, 0.14] |
| Proportion of people working in the third industry (median [IQR]) | 0.30 [0.29, 0.32] |
| Taxable income per person (1,000 yens) (median [IQR]) | 1191.23 [1013.90, 1357.37] |
| Basic features |  |
| Population (100,000 persons) (median [IQR]) | 2.24 [1.04, 4.77] |
| Area (km<sup>2</sup>) (median [IQR]) | 855.66 [434.46, 1405.56] |
| Proportion of habitable area (median [IQR]) | 0.37 [0.25, 0.56] |
| Population density (/ha) (median [IQR]) | 2.46 [0.94, 6.90] |
| RAM (median [IQR]) | 1.03 [0.95, 1.13] |

SMA, secondary medical area; IQR, Interquartile range; ePCI, emergency percutaneous coronary intervention; y/o, years old; AMI, acute myocardial infarction; RAM, risk adjusted mortality.

Figure 1 shows the sorted bar plot of the SMAs’ RAM. There is wide variation in RAM; the maximum value was 0.593 and the minimum value was 1.445. The ratio of the maximum to the minimum was 2.436.

**Figure 1.**
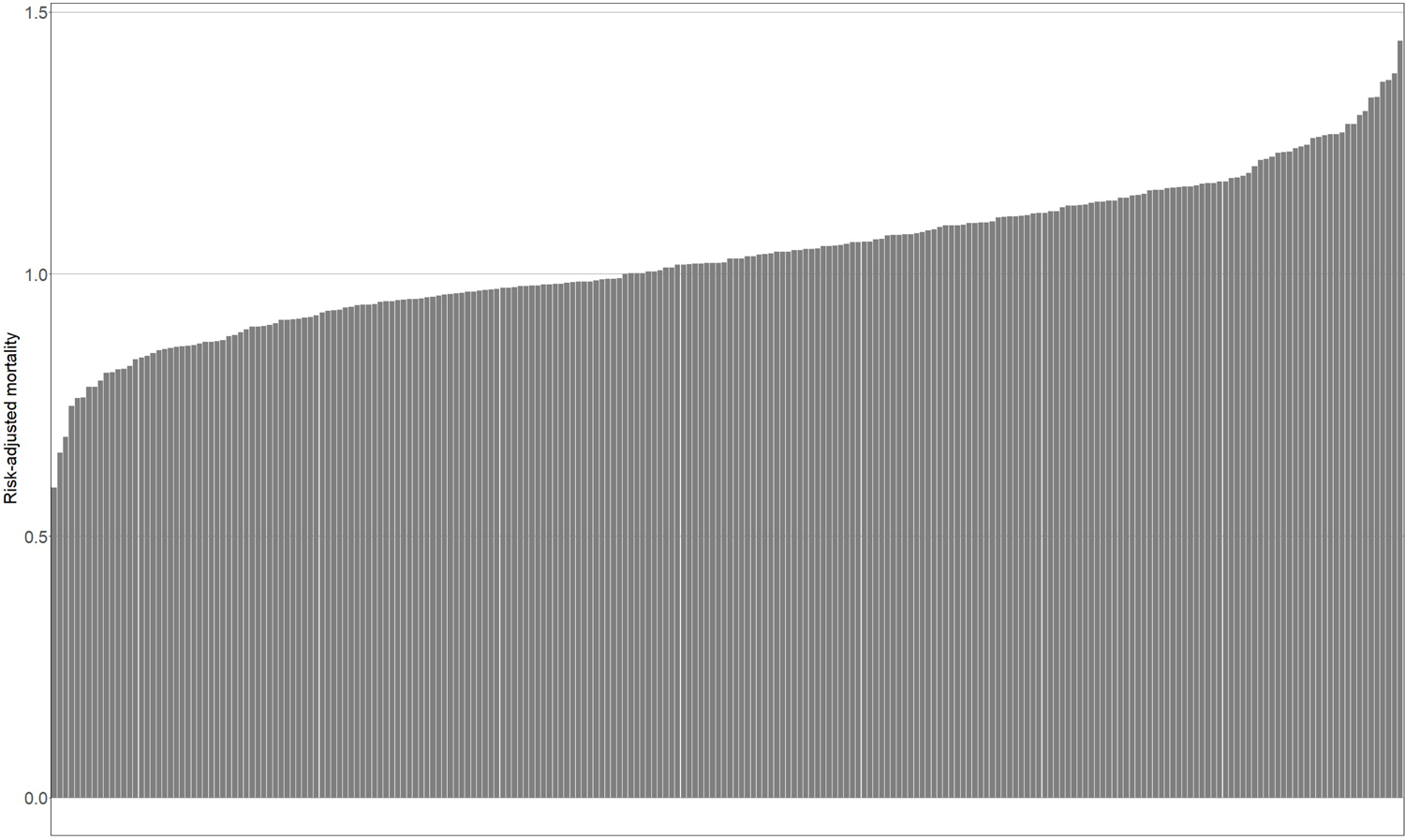
Risk-Adjusted Mortality (RAM) of Secondary Medical Areas (SMAs) The RAM of SMAs is plotted in ascending order.

### SMAs’ AMI Case Volumes and Crude Mortality

Supplementary Figure 1 plots the SMAs’ AMI case volumes and crude mortality. There is a strong negative correlation between case volumes and crude mortality (correlation coefficient: −0.412; 95% confidence interval: −0.467 to −0.353), justifying the imputation of crude mortality based on the distribution of case volume.

### Regression Analyses

OLS regression gave large Variance Inflation Factors (VIFs) for some variables. Specifically, 50.7 for the proportion working in the second industry, 48.8 for the number of hospitals per area, and 39.5 for the proportion working in the first industry. These large VIFs demonstrate strong multicollinearity in the data.

With the PLS regression, the cumulative explained variances of the dependent variable by components 1 to 5 were 0.1765, 0.2137, 0.2223, 0.2246, and 0.2269. More than two components could create a minor difference to the explained variance, and therefore, we chose two as the number of components.

Figures 2–5 and Table 2 show the results of the PLS regression. Figures 2 and 3 show the plot of the SMAs’ RAM with the scores of Component 1 and with those of component 2. There were positive correlations between RAM and the scores of Components 1 and 2. Since low RAM indicates good quality of AMI care, small scores of Components 1 and 2 were supposed to correlate with good quality.

**Figure 2.**
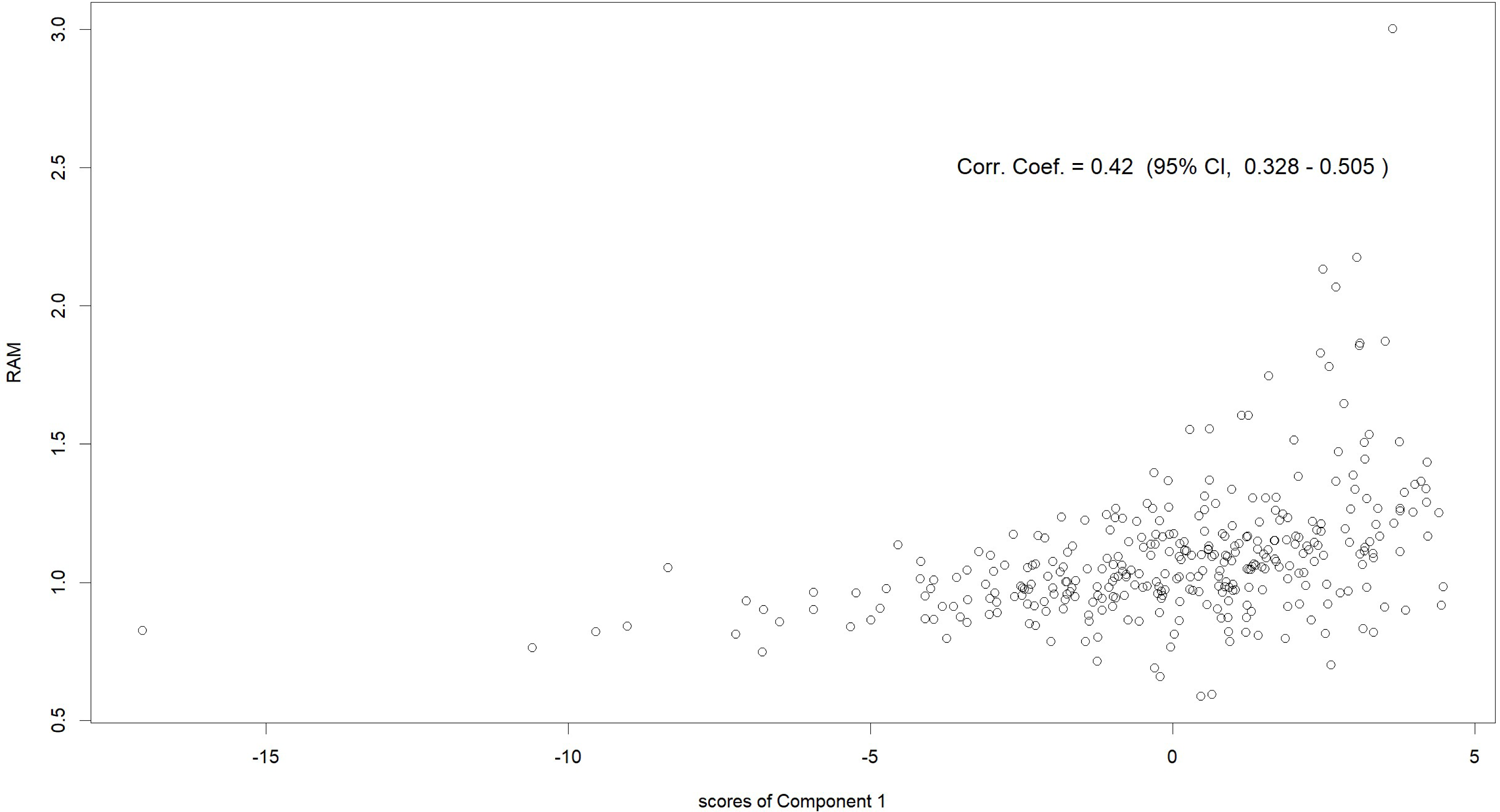
Component 1 Scores and RAM of SMAs Scores of Component 1 in the horizontal axis and RAM in the vertical axis. SMA, secondary medical area; RAM, risk-adjusted mortality; corr., correlation; coef., coefficient; CI, confidence interval.

**Figure 3.**
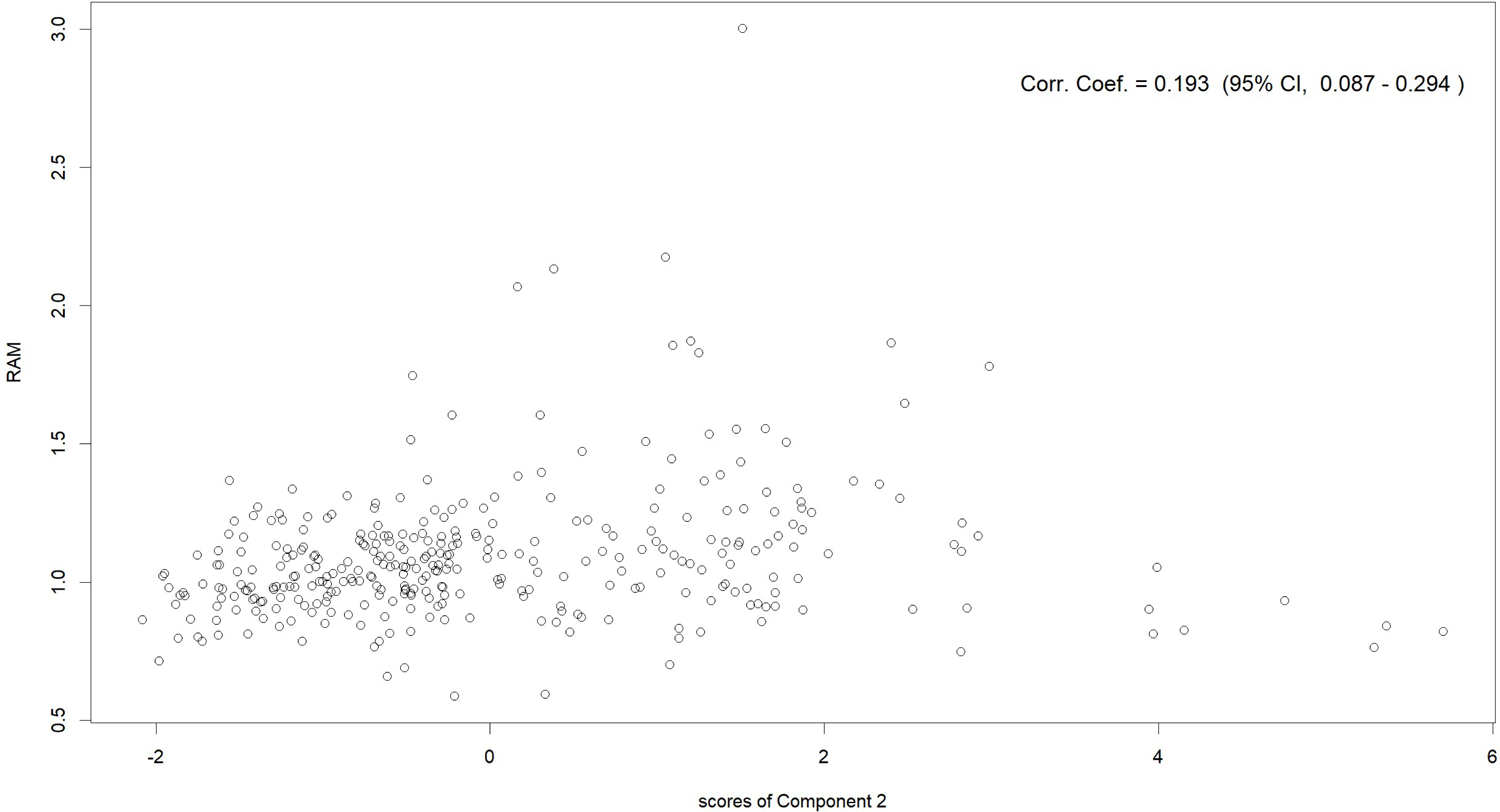
Component 2 Scores and RAM of SMAs Scores of Component 2 in the horizontal axis, and RAM in the vertical axis. SMA, secondary medical area; RAM, risk-adjusted mortality; corr., correlation; coef., coefficient; CI, confidence interval.

**Figure 4.**
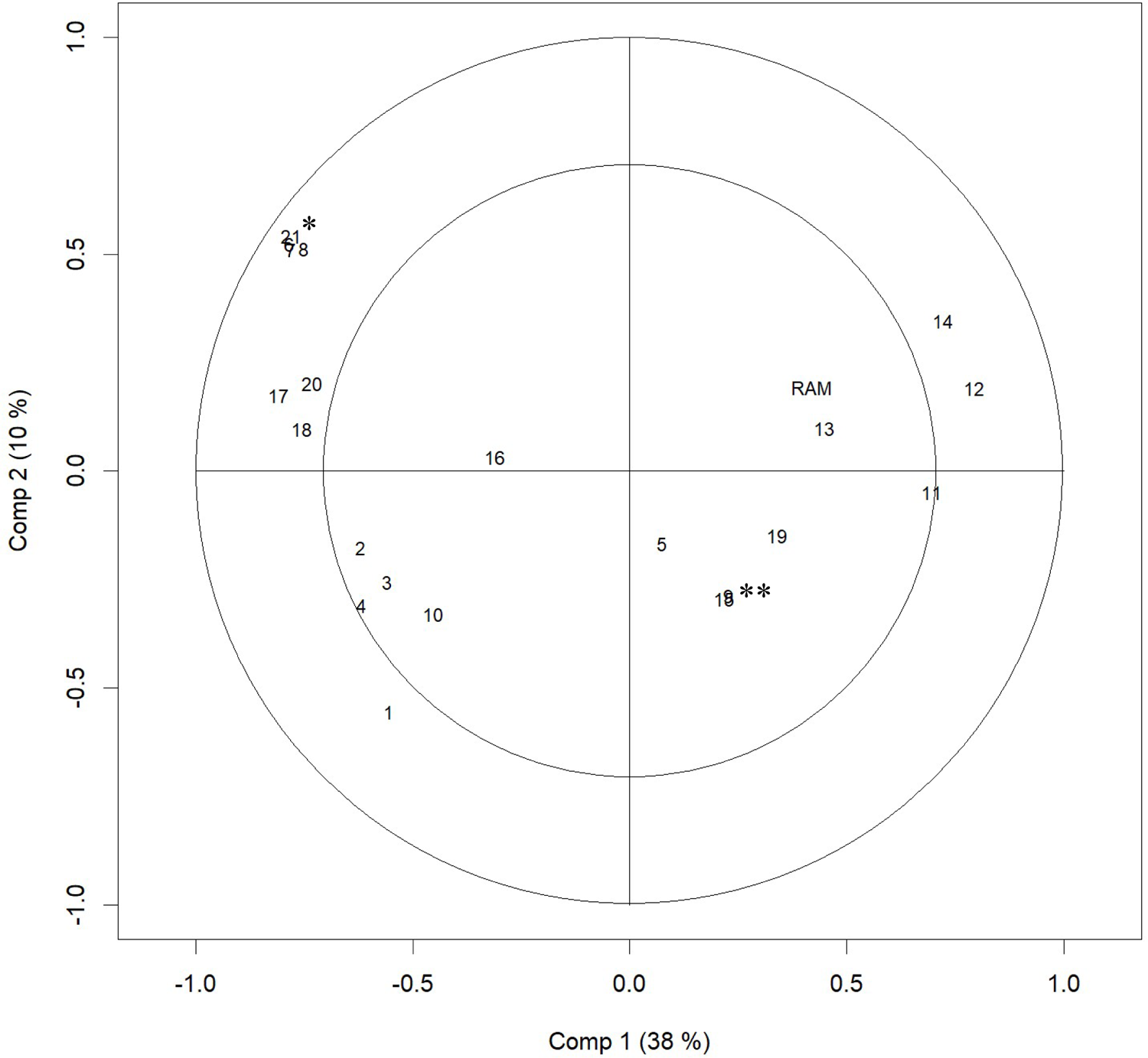
Correlation Loading Plot of PLS Regression Results The plot of numbers indicates the loadings of variables to Component 1 and 2. The plot of RAM indicates its correlation with Components 1 and 2. The distance from the origin indicates the variance of the variable explained by Components 1 and 2. The inner and the outer circles indicate 50% and 100% explained variance, respectively. Each number indicates as following aspects: 1, the Indicator of Access; 2, number of all physicians per resident; 3, number of cardiologists per resident; 4, number of cardiovascular surgeons per resident; 5, number of beds per resident; 6, number of emergency hospitals per area; 7, number of hospitals per area; 8, number of clinics per area; 9, medical expenditure per person; 10, population proportion, under 14 y/o; 11, population proportion, 65–74 y/o; 12, population proportion, over 75 y/o; 13, proportion of people working; 14, proportion of people working in the first industry; 15, proportion of people working in the second industry; 16, proportion of people working in the third industry; 17, taxable income per person; 18, population; 19, area; 20, proportion of habitable area; 21, population density. * indicates numbers of 6, 7, 8, and 21 ** indicates numbers of 9 and 15 PLS, partial least squares; ePCI, emergency percutaneous coronary intervention; y/o, years old; RAM, risk-adjusted mortality.

**Figure 5.**
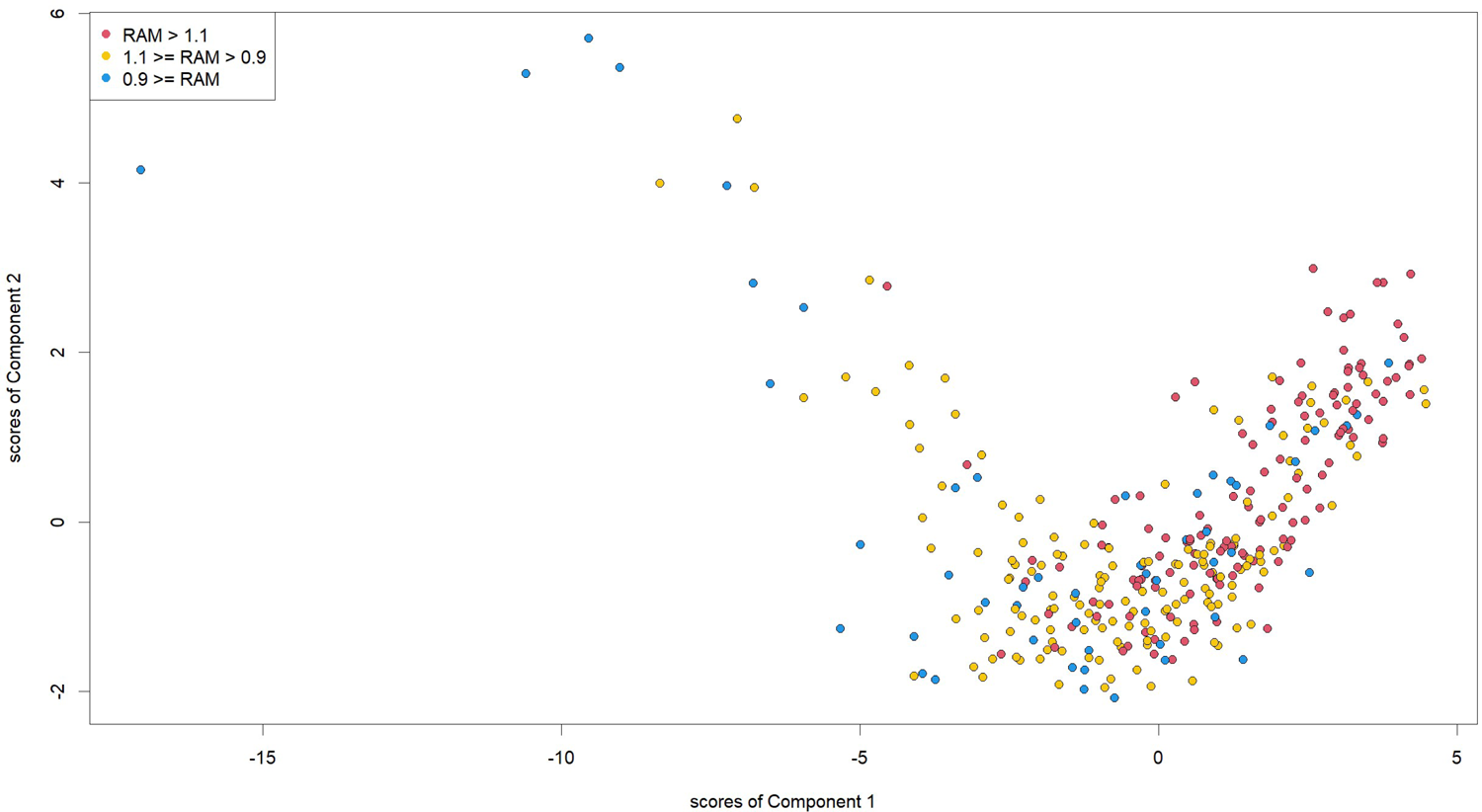
SMA Scores for Components 1 and 2 Points indicate SMAs’ scores of Components 1 and 2. Red points indicate SMAs where RAM is over 1.1. Green points indicate SMAs where RAM is 0.9–1.1. Blue points indicate SMAs where RAM is equal to or under 0.9. PLS, partial least squares; RAM, risk-adjusted mortality; SMA, secondary medical area.

**Table 2.**
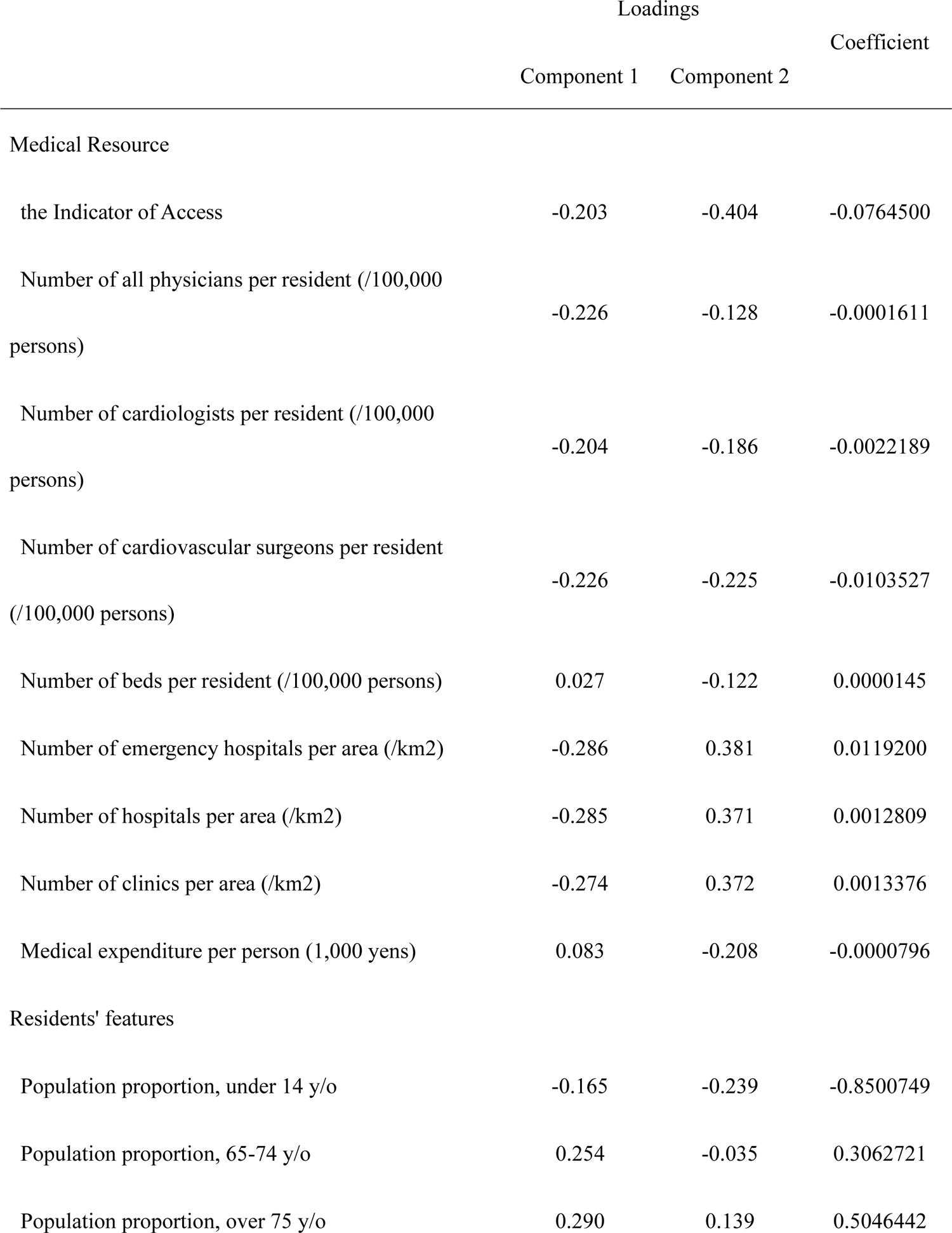
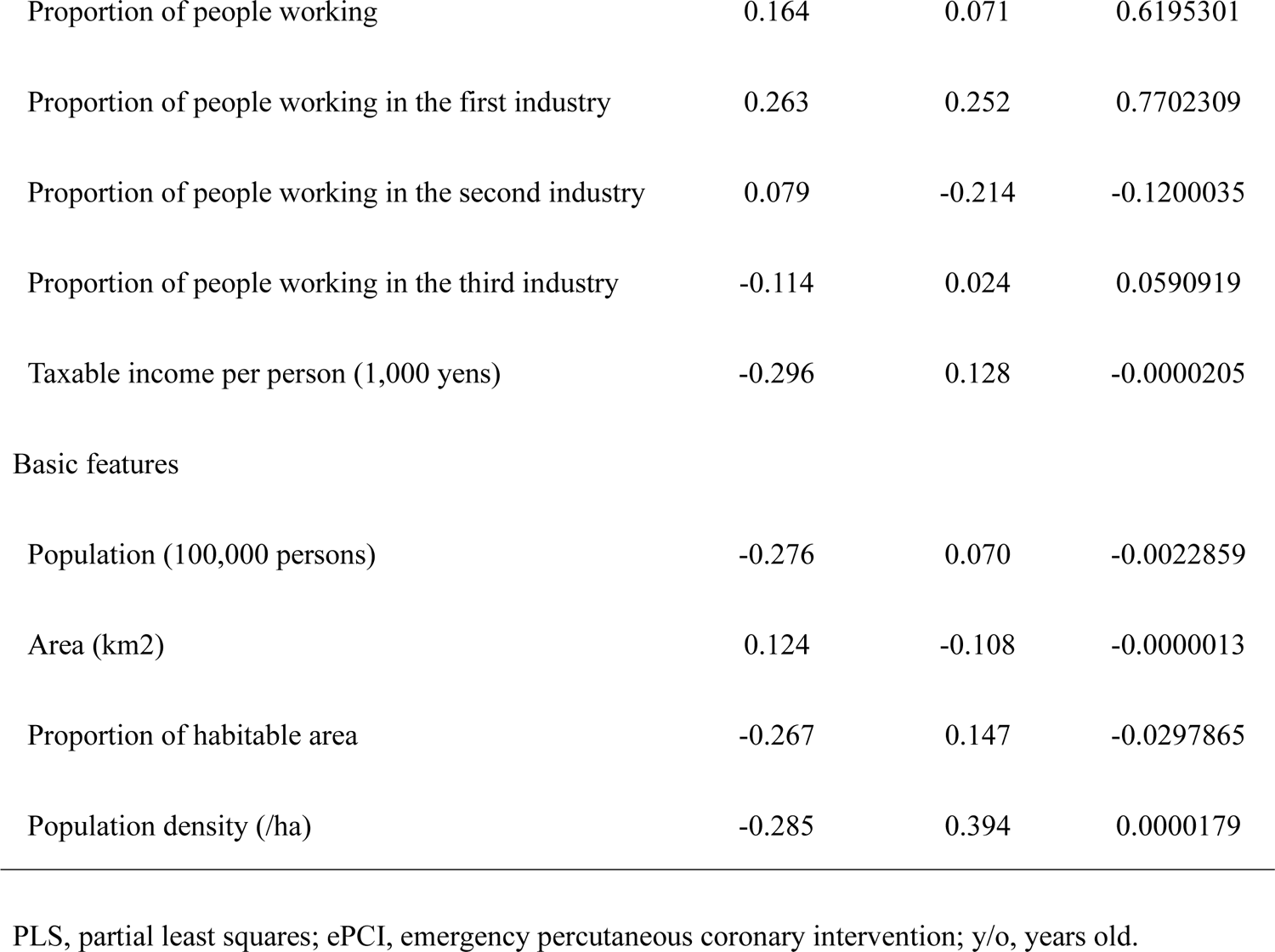
Loadings and coefficient of the regional variables from the PLS regression analysis

Figure 4 and Table 2 show each variable’s coefficient and loadings with Components 1 and 2. With Component 1, the population proportion of the elderly (65– 74 y/o and 75+ y/o), and the proportion working in the first industry had a positive loading, and the number of medical facilities (emergency hospitals, hospitals, and clinics) per area, the population density, the taxable income per persons, the population, and the proportion of habitable area had a negative loading. With Component 2, the numbers of medical facilities (emergency hospitals, hospitals, and clinics) per area and the population density had a positive loading, while the Indicator of Access and the number of physicians (i.e., the number of all physicians, cardiologists, and cardiovascular surgeons) per person had a negative loading.

Figure 5 plots SMAs’ scores of Components 1 and 2. SMAs in which the scores of Component 1 were close to the smaller and larger ends show relatively large scores for Component 2.

Supplementary Tables 2 to 5 show the loadings and the coefficients resulting from sensitivity analysis. There was no substantial difference with the main analysis.

## DISCUSSION

While we adjust regional variation of risk of patients at admission, there was substantial regional variation in the mortality of AMI patients. There was a 2.436 times difference in the risk-adjusted in-hospital mortality. Through PLS regression analysis, we identified two components that influence the regional quality of healthcare for AMI patients, as follows.

Component 1 can be interpreted as the degree of rurality, given the features of having a large elderly proportion,[30] low population density, low income,[31] and poor medical human resources[32–34] match the characteristics of rural areas in Japan. The suggested negative influence of rurality on healthcare quality is consistent with previous findings.[13] In addition, having sufficient medical resources was suggested to improve healthcare quality for AMI patients at a hospital level,[15] which was consistent with our regional-level investigation. The proportion of the elderly, whose loading for Component 1 was positive, can be interpreted as one aspect of the insufficiency of medical resources. Large proportions of the elderly mean a large demand for healthcare, resulting in more burden on the system and a scarcity of medical resources.

Component 2 positively correlated with the population density and the number of medical facilities per area, but negatively correlated with the number of physicians per person and the Indicator of Access, which cannot be explained by the degree of rurality. One possible explanation of Component 2 is that it reflects not only the degree of richness of medical resources but also the integration of healthcare delivery– coordinated healthcare delivery. Less access to high-volume ePCI centres and more medical facilities per area possibly mean that healthcare provision is fragmented and less integrated, meaning healthcare delivery is not well-coordinated. Although the observational evidence at a regional level was scarce, fragmentation of the healthcare system has been theoretically suggested to reduce performance.[35]

Loadings of the variables can explain the score plot of the SMAs. When the population density and number of facilities per area are large, the score of Component 1 tends to be small, but the score of Component 2 tends to be large. In fact, SMAs in which the scores of Component 1 were close to the lower end show relatively high scores in Component 2. In contrast, when the two indicators of Access and physicians per person are low, the scores of Component 1 and 2 tend to be high. Indeed, SMAs in which the scores of Component 2 were close to the higher end show relatively high scores for Component 2.

Policy actions are needed to address the unwarranted disparity in healthcare quality; the discussion above suggests that the approaches required to improve quality are different between rural and urban areas. In rural areas, the required policy actions should target the reduction of Components 1 and 2 simultaneously; the score plot shows the positive correlation between Components 1 and 2 for SMAs with high scores for Component 1. For example, medical resources are considered to be sensitive to health policies and increasing medical resources can reduce the scores in both Components 1 and 2. Uneven distribution of medical professionals is recognised as an important issue in Japanese health policy and our results add further rationale to solving regional disparities. At the same time, increasing the Indicator of Access can also decrease the scores of Components 1 and 2.

In contrast, in urban areas, the importance of utilizing medical resources—a part of Component 2—should be stressed more. From the perspective of addressing the uneven distribution of medical resources, policies to decrease the scores of Component 1 in urban areas is difficult to justify. However, there is the possibility of increasing efficiency, including increasing the Indicator of Access; the score plot shows that a high percentage of SMAs with low scores for Component 1 had high scores for Component 2. However, it should be noted that compared with Component 1, the explained variance in RAM by Component 2 was small, meaning the effect of reducing fragmentation is less significant than decreasing rurality.

The loadings of the variables, shown in Figure 5, highlight the importance of access to high-volume ePCI centres. That indicator had negative loadings both for Components 1 and 2, and the distance from the origin was large, meaning there was a large correlation with Components 1 and 2. This result suggests the potential of increasing access to high-volume ePCI centres, to improve healthcare quality. In fact, the hub-and-spoke model—where a network between centres providing resource-intensive services (i.e., the spokes) and facilities providing more primary services (i.e., the hub) is established—has been posited as an effective way to improve outcomes,[36, 37] which is consistent with our results.

This study has several limitations. First, although the DPC data for the majority of patients who received acute case treatment was included in the MHLW DPC database,[20] the calculation of each SMA’s RAM for the patients whose DPC data were available could introduce selection bias. Second, as described, there were missing RAM values due to data availability. We performed imputation as justified by the data, and sensitivity analyses showed that the conclusion of the study was valid, but the potential bias due to the missing values cannot be denied. Third, this study did not consider the many unobservable factors that can influence healthcare quality. For instance, the education level of health professionals, on which data are not available at the regional level, can improve healthcare quality. In addition, the results implied the fragmentation of healthcare provision might influence the quality, but many factors other than those involved in this study can also be associated with the fragmentation. Further research is warranted to clarify the mechanism that produces good quality of healthcare at a regional level.

## CONCLUSION

There was wide regional variation in the quality of in-hospital healthcare for AMI patients. Results suggest that, at a regional level, the degree of rurality, the sufficiency of medical resources, access to high-volume ePCI centres, and coordination of healthcare delivery were associated with healthcare quality for AMI patients.

## Data Availability

Data cannot be shared for ethical/privacy reasons.

## DECRALATION

### ETHICS APPROVAL AND CONSENT TO PARTICIPATE

This study was conducted in accordance with the principles of the Declaration of Helsinki and the study was approved by the Ethics Committee, Kyoto University Graduate School and Faculty of Medicine (approval numbers: R0135, R1389, and R2062) with a waiver of informed consent prior to data collection.

## CONSENT FOR PUBLICATION

Not applicable

### AVAILABILITY OF DATA AND MATERIALS

Data cannot be shared for ethical/privacy reasons.

## AUTHORS’ CONTRIBUTIONS

S.W., J.S., S.K., and Y.I. were involved in the conceptualization of the study and the design of the methodology. J.S. was in charge of the data curation. S.W. and J.S. undertook the formal analysis. S.W. wrote the original draft of the manuscript with input from all authors. All authors critically revised the report, commented on drafts of the manuscript, and approved the final report. Y.I. was in charge of the administration and supervision of the project administration.

---

**Supplementary Figure 1.**
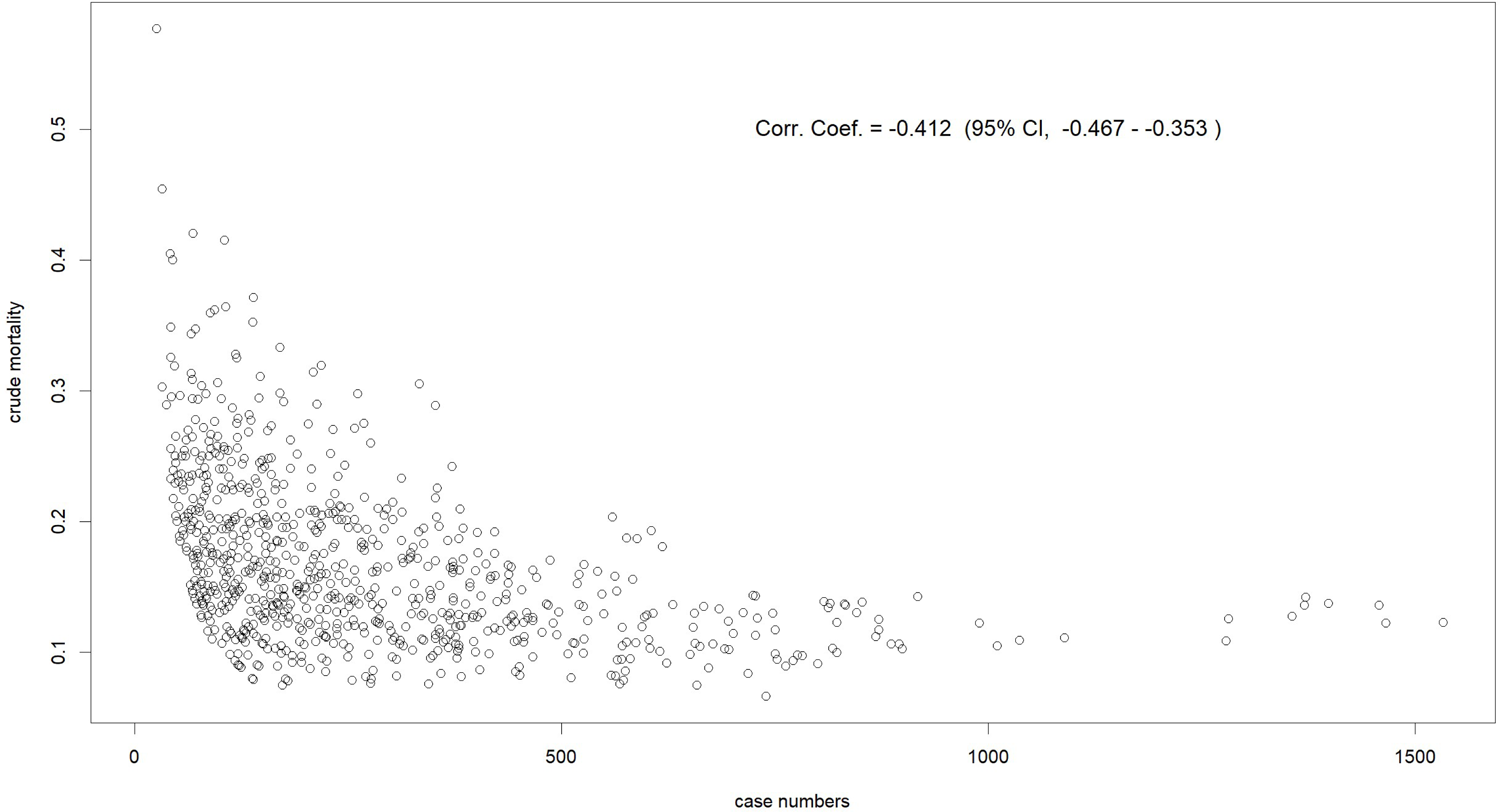
SMAs’ AMI Case Numbers and Crude Mortality SMA, secondary medical area; AMI, acute myocardial infarction.

**Supplementary Table 1.** Reference of regional variables

|  | Fiscal Year | Reference |
| --- | --- | --- |
| Medical Resource |  |  |
| the Indicator of Access | 2016-2018 | National Database of Health Insurance Claims and Specific Health Checkups of Japan * |
| Number of all physicians per resident<br>(/100,000 persons) | 2016 | Survey of Physicians, Dentists, and Pharmacists (MHLW) |
| Number of cardiologists per resident<br>(/100,000 persons) | 2016 | Survey of Physicians, Dentists, and Pharmacists (MHLW) |
| Number of cardiovascular surgeons per resident<br>(/100,000 persons) | 2016 | Survey of Physicians, Dentists, and Pharmacists (MHLW) |
| Number of beds per resident<br>(/100,000 persons) | 2016 | Survey of Medical Facilities (MHLW) |
| Number of emergency hospitals per area (/km2) | 2017 | Public data of the reports of the beds functions (MHLW) |
| Number of hospitals per area (/km2) | 2016 | Survey of Medical Facilities (MHLW) |
| Number of clinics per area (/km2) | 2016 | Survey of Medical Facilities (MHLW) |
| Medical expenditure per person (1,000 yens) | 2018 | Regional difference of the medical expenditure (MHLW) |
| Residents' features |  |  |
| Population proportion, under 14 y/o | 2017 | Population, demographics, and the number of households<br>from Basic Resident Register (MIAC) |
| Population proportion, 65-74 y/o | 2017 | Population, demographics, and the number of households<br>from Basic Resident Register (MIAC) |
| Population proportion, over 75 y/o | 2017 | Population, demographics, and the number of households<br>from Basic Resident Register (MIAC) |
| Proportion of people working | 2015 | National Census (MIAC) |
| Proportion of people working in the first industry | 2015 | National census (MIAC) |
| Proportion of people working in the second industry | 2015 | National census (MIAC) |
| Proportion of people working in the third industry | 2015 | National census (MIAC) |
| Taxable income per person (1,000 yens) | 2015 | Survey of taxes of shi, ku, machi, mura (MIAC) |
| Basic features |  |  |
| Population (100,000 persons) | 2017 | Population, demographics, and the number of households<br>from Basic Resident Register (MIAC) |
| Area (km2) | 2017 | Survey of area of shi, ku, machi, mura (MIAC) |
| Proportion of habitable area | 2017 | Survey of area of shi, ku, machi, mura (MIAC) |
| Population density (/ha) | 2017 | Survey of taxes of shi, ku, machi, mura (MIAC),<br>Population, demographics, and the number of households<br>from Basic Resident Register (MIAC) |
| Quality of AMI healthcare |  |  |
| RAM (median [IQR]) | 2016-2018 | -* |

ePCI, emergency percutaneous coronary intervention; y/o, years old; MHLW, Ministry of Health, Labour, and Welfare; MIAC, Ministry of Internal Affairs and Communication
\* the way of calculation was described in the method section.

**Supplementary Table 2.** Loadings and coefficient of the regional variables from the PLS regression analysis of the first sensitivity analysis for the imputation for missing values of observed deaths (the crude mortality proportion used for the estimation was the proportion in SMAs where case volumes were under the maximum value of the case volumes of SMAs with missing values)

|  | Loadings |  | Coefficient |
| --- | --- | --- | --- |
|  | Component 1 | Component 2 |  |
| Medical Resource |  |  |  |
| the Indicator of Access | -0.192 | -0.343 | -0.0414727 |
| Number of all physicians per resident (/100,000 persons) | -0.229 | -0.226 | -0.0001314 |
| Number of cardiologists per resident (/100,000 persons) | -0.204 | -0.270 | -0.0018010 |
| Number of cardiovascular surgeons per resident (/100,000 persons) | -0.224 | -0.278 | -0.0076604 |
| Number of beds per resident (/100,000 persons) | 0.019 | -0.242 | -0.0000008 |
| Number of emergency hospitals per area (/km2) | -0.289 | 0.344 | -0.0188681 |
| Number of hospitals per area (/km2) | -0.288 | 0.331 | -0.0156930 |
| Number of clinics per area (/km2) | -0.278 | 0.327 | 0.0003380 |
| Medical expenditure per person (1,000 yens) | 0.077 | -0.317 | -0.0000678 |
| Residents' features |  |  |  |
| Population proportion, under 14 y/o | -0.155 | -0.178 | -0.5643065 |
| Population proportion, 65-74 y/o | 0.246 | -0.095 | 0.1957692 |
| Population proportion, over 75 y/o | 0.278 | 0.068 | 0.3064798 |
| Proportion of people working | 0.169 | 0.191 | 0.5427202 |
| Proportion of people working in the first industry | 0.255 | 0.240 | 0.5596337 |
| Proportion of people working in the second industry | 0.092 | -0.055 | 0.0265096 |
| Proportion of people working in the third industry | -0.117 | -0.059 | -0.0926922 |
| Taxable income per person (1,000 yens) | -0.291 | 0.152 | -0.0000170 |
| Basic features |  |  |  |
| Population (100,000 persons) | -0.273 | 0.064 | -0.0016908 |
| Area (km<sup>2</sup>) | 0.124 | -0.088 | 0.0000010 |
| Proportion of habitable area | -0.263 | 0.176 | -0.0198652 |
| Population density (/ha) | -0.287 | 0.370 | -0.0000603 |

PLS, partial least squares; SMA, secondary medical area; PLS, partially least squares; ePCI, emergency percutaneous coronary intervention; y/o, years old.

**Supplementary Table 3.** Loadings and coefficient of the regional variables from the PLS regression analysis of the second sensitivity analysis for the imputation for missing values of observed deaths (limiting to the SMAs where exact observed deaths were obtained)

|  | Loadings |  | Coefficient |
| --- | --- | --- | --- |
|  | Component 1 | Component 2 |  |
| Medical Resource |  |  |  |
| the Indicator of Access | -0.120 | -0.311 | -0.0381312 |
| Number of all physicians per resident (/100,000 persons) | -0.203 | -0.191 | -0.0000651 |
| Number of cardiologists per resident (/100,000 persons) | -0.192 | -0.282 | -0.0020271 |
| Number of cardiovascular surgeons per resident (/100,000 persons) | -0.188 | -0.222 | -0.0037028 |
| Number of beds per resident (/100,000 persons) | 0.026 | -0.194 | -0.0000001 |
| Number of emergency hospitals per area (/km2) | -0.306 | 0.312 | -0.0283548 |
| Number of hospitals per area (/km2) | -0.304 | 0.299 | -0.0197228 |
| Number of clinics per area (/km2) | -0.299 | 0.293 | -0.0002248 |
| Medical expenditure per person (1,000 yens) | 0.081 | -0.384 | -0.0001071 |
| Residents' features |  |  |  |
| Population proportion, under 14 y/o | -0.100 | -0.377 | -0.7499074 |
| Population proportion, 65-74 y/o | 0.250 | -0.008 | 0.3584526 |
| Population proportion, over 75 y/o | 0.268 | 0.120 | 0.3769717 |
| Proportion of people working | 0.196 | 0.172 | 0.3997076 |
| Proportion of people working in the first industry | 0.248 | 0.241 | 0.5406638 |
| Proportion of people working in the second industry | 0.159 | -0.022 | 0.0767382 |
| Proportion of people working in the third industry | -0.110 | -0.078 | -0.1163348 |
| Taxable income per person (1,000 yens) | -0.299 | 0.139 | -0.0000226 |
| Basic features |  |  |  |
| Population (100,000 persons) | -0.259 | 0.070 | -0.0010969 |
| Area (km2) | 0.152 | 0.024 | 0.0000052 |
| Proportion of habitable area | -0.253 | 0.172 | -0.0173603 |
| Population density (/ha) | -0.306 | 0.330 | -0.0000870 |

PLS, partial least squares; SMA, secondary medical area; PLS, partially least squares; ePCI, emergency percutaneous coronary intervention; y/o, years old.

**Supplementary Table 4.** Loadings and coefficient of the regional variables from the PLS regression analysis of the first sensitivity analysis for the ePCI-large-hospitals share (SMAs where no ePCIs were performed were removed)

|  | Loadings |  | Coefficient |
| --- | --- | --- | --- |
|  | Component 1 | Component 2 |  |
| Medical Resource |  |  |  |
| the Indicator of Access | -0.160 | -0.376 | -0.0557234 |
| Number of all physicians per resident (/100,000 persons) | -0.221 | -0.206 | -0.0001225 |
| Number of cardiologists per resident (/100,000 persons) | -0.194 | -0.253 | -0.0021187 |
| Number of cardiovascular surgeons per resident (/100,000 persons) | -0.214 | -0.263 | -0.0086165 |
| Number of beds per resident (/100,000 persons) | 0.036 | -0.185 | 0.0000270 |
| Number of emergency hospitals per area (/km2) | -0.304 | 0.317 | -0.0070148 |
| Number of hospitals per area (/km2) | -0.303 | 0.304 | -0.0094333 |
| Number of clinics per area (/km2) | -0.294 | 0.302 | 0.0006723 |
| Medical expenditure per person (1,000 yens) | 0.071 | -0.377 | -0.0001107 |
| Residents' features |  |  |  |
| Population proportion, under 14 y/o | -0.123 | -0.005 | -0.0691534 |
| Population proportion, 65-74 y/o | 0.234 | -0.265 | -0.2726920 |
| Population proportion, over 75 y/o | 0.271 | -0.017 | 0.3106837 |
| Proportion of people working | 0.178 | 0.250 | 0.7138245 |
| Proportion of people working in the first industry | 0.265 | 0.334 | 0.9101322 |
| Proportion of people working in the second industry | 0.106 | -0.045 | -0.0055128 |
| Proportion of people working in the third industry | -0.122 | -0.107 | -0.2078844 |
| Taxable income per person (1,000 yens) | -0.299 | 0.117 | -0.0000274 |
| Basic features |  |  |  |
| Population (100,000 persons) | -0.277 | 0.037 | -0.0018993 |
| Area (km<sup>2</sup>) | 0.142 | -0.045 | 0.0000015 |
| Proportion of habitable area | -0.266 | 0.209 | -0.0127545 |
| Population density (/ha) | -0.303 | 0.340 | -0.0000343 |

SMA, secondary medical area; PLS, partially least squares; ePCI, emergency percutaneous coronary
intervention; y/o, years old.

**Supplementary Table 5.** Loadings and coefficient of the regional variables from the PLS regression analysis of the second sensitivity analysis for the ePCI-large-hospitals share (the indicator was set to one if there is only one hospital which undertook ePCIs in a SMA even when case volumes of those hospitals were under the threshold)

|  | Loadings |  | Coefficient |
| --- | --- | --- | --- |
|  | Component 1 | Component 2 |  |
| Medical Resource |  |  |  |
| the Indicator of Access | -0.131 | -0.407 | -0.0972094 |
| Number of all physicians per resident (/100,000 persons) | -0.230 | -0.138 | -0.0001738 |
| Number of cardiologists per resident (/100,000 persons) | -0.206 | -0.202 | -0.0024081 |
| Number of cardiovascular surgeons per resident (/100,000 persons) | -0.226 | -0.232 | -0.0113941 |
| Number of beds per resident (/100,000 persons) | 0.026 | -0.135 | 0.0000169 |
| Number of emergency hospitals per area (/km2) | -0.294 | 0.379 | 0.0212653 |
| Number of hospitals per area (/km2) | -0.292 | 0.370 | 0.0059918 |
| Number of clinics per area (/km2) | -0.282 | 0.368 | 0.0016557 |
| Medical expenditure per person (1,000 yens) | 0.085 | -0.201 | -0.0000870 |
| Residents' features |  |  |  |
| Population proportion, under 14 y/o | -0.166 | -0.258 | -0.9340170 |
| Population proportion, 65-74 y/o | 0.258 | -0.019 | 0.3258465 |
| Population proportion, over 75 y/o | 0.292 | 0.159 | 0.5529598 |
| Proportion of people working | 0.167 | 0.077 | 0.6598186 |
| Proportion of people working in the first industry | 0.261 | 0.243 | 0.8615148 |
| Proportion of people working in the second industry | 0.086 | -0.204 | -0.1541198 |
| Proportion of people working in the third industry | -0.115 | 0.029 | 0.0574371 |
| Taxable income per person (1,000 yens) | -0.300 | 0.122 | -0.0000222 |
| Basic features |  |  |  |
| Population (100,000 persons) | -0.279 | 0.068 | -0.0024803 |
| Area (km<sup>2</sup>) | 0.127 | -0.109 | -0.0000016 |
| Proportion of habitable area | -0.272 | 0.141 | -0.0317202 |
| Population density (/ha) | -0.292 | 0.392 | 0.0000389 |

PLS, partial least squares; SMA, secondary medical area; PLS, partially least squares; ePCI, emergency percutaneous coronary intervention; y/o, years old.

## REFERENCES

1. Sakamoto H, Rahman M, Nomura S, et al. Japan Health System Review. World Health Organization, Regional Office for South-East Asia. 2018.

2. Ministry of Health, Labour, and Welfare, Japan. Regional Medical Plan. https://www.mhlw.go.jp/stf/seisakunitsuite/bunya/kenkou_iryou/iryou/iryou_keikaku/index.html (accessed 1 Mar 2022).

3. Sugahara T. Economic background and issues of regional medical coordination policy in Japan (Strategic management of evidence-based health and medical care policy: How to use new Digital Big Data in health care system). J Natl Inst Public Heal 2013;62:36–45.

4. Cabinet Office Japan. Annual Report on the Ageing Society [Summary] FY2020. 2020.https://www8.cao.go.jp/kourei/english/annualreport/2020/pdf/2020.pdf (accessed 26 Feb 2022).

5. Ministry of Health, Labour, and Welfare, Japan. Concept of regional health systems (‘Chiiki Iryo Koso’ in Japanese). https://www.mhlw.go.jp/stf/seisakunitsuite/bunya/0000080850.html (accessed 26 Feb 2022).

6. National commission for the reform of the social security system. Report of National commission for the reform of the social security system. 2013. https://www.kantei.go.jp/jp/singi/kokuminkaigi/index.html (accessed 26 Feb 2022).

7. Lohr KN, Committee to Design a Strategy for Quality Review and Assurance in Medicare I of M. Medicare: a strategy for quality assurance, Volume 1. Washington D.C. NATIONAL ACADEMY PRESS. 1990.

8. Petersen LA, Woodard LD, Urech T, et al. Annals of Internal Medicine Improving Patient Care Does Pay-for-Performance Improve the Quality of Health Care ?. Ann Intern Med 2006;145:265–72.

9. Shine D. Risk-adjusted mortality: Problems and possibilities. Comput Math Methods Med;2012.

10. O’Connor GT, Quinton HB, Traven ND, et al. Geographic variation in the treatment of acute myocardial infarction: The cooperative cardiovascular project. J Am Med Assoc 1999;281:627–33.

11. Kim J, Yang KH, Choi AR, et al. Healthcare quality assessments: No guarantees of same outcomes for different socio-economic stroke patients. Int J Qual Heal Care 2021;33:1–6.

12. Ko DT, Krumholz HM, Wang Y, et al. Regional differences in process of care and outcomes for older acute myocardial infarction patients in the United States and Ontario, Canada. Circulation 2007;115:196–203.

13. Kim HS, Kang DR, Kim I, et al. Comparison between urban and rural mortality in patients with acute myocardial infarction: A nationwide longitudinal cohort study in South Korea. BMJ Open 2020;10:1–6.

14. Pross C, Busse R, Geissler A. Hospital quality variation matters – A time-trend and cross-section analysis of outcomes in German hospitals from 2006 to 2014. Health Policy 2017;121:842–52.

15. Park S, Lee J, Ikai H, et al. Decentralization and centralization of healthcare resources: Investigating the associations of hospital competition and number of cardiologists per hospital with mortality and resource utilization in Japan. Health Policy 2013;113:100–9.

16. Shen Y-C, Hsia RY. Association Between Ambulance Diversion And Survival Among Patients with Acute Myocardial Infarction. JAMA 2011;305:2440–7.

17. Fairfield KM, Black AW, Lucas FL, et al. Behavioral Risk Factors and Regional Variation in Cardiovascular Health Care and Death. Am J Prev Med 2018;54:376–84.

18. Tsugawa Y, Hasegawa K, Hiraide A, et al. Regional health expenditure and health outcomes after out-of-hospital cardiac arrest in Japan: An observational study. BMJ Open 2015;5:1–10.

19. Hosokawa R, Ojima T, Myojin T, et al. Associations between healthcare resources and healthy life expectancy: A descriptive study across secondary medical areas in Japan. Int J Environ Res Public Health 2020;17:1–16.

20. Hayashida K, Murakami G, Matsuda S, et al. History and profile of diagnosis procedure combination (DPC): Development of a real data collection system for acute inpatient care in Japan. J Epidemiol 2021;31:1–11.

21. Otsubo T, Goto E, Morishima T, et al. Regional variations in in-hospital mortality, care processes, and spending in acute ischemic stroke patients in Japan. J Stroke Cerebrovasc Dis 2015;24:239–51.

22. Genta KATO. History of the Secondary Use of National Database of Health Insurance Claims and Specific Health Checkups of Japan(NDB). Trans Japanese Soc Med Biol Eng 2017;55:143–50.

23. Mohammed MA, Manktelow BN, Hofer TP. Comparison of four methods for deriving hospital standardised mortality ratios from a single hierarchical logistic regression model. Stat Methods Med Res 2016;25:706–15.

24. Badhiwala JH, Khan O, Wegner A, et al. A partial least squares analysis of functional status, disability, and quality of life after surgical decompression for degenerative cervical myelopathy. Sci Rep 2020;10:1–11.

25. Ibrahim GM, Morgan BR, Fallah A. A partial least squares analysis of seizure outcomes following resective surgery for tuberous sclerosis complex in children with intractable epilepsy. Child’s Nerv Syst 2015;31:181–4.

26. Young JM, Morgan BR, Mišić B, et al. A Partial Least-Squares Analysis of Health-Related Quality-of-Life Outcomes After Aneurysmal Subarachnoid Hemorrhage. Neurosurgery 2015;77:908–15.

27. Wimberly MC, Lamsal A, Giacomo P, et al. Regional variation of climatic influences on West Nile virus outbreaks in the United States. Am J Trop Med Hyg 2014;91:677–84.

28. Wang H, Leung GM, Mary Schooling C. Life course body mass index and adolescent self-esteem: Evidence from Hong Kong’s ‘children of 1997’ Birth Cohort. Obesity 2015;23:429–35.

29. Liland KH, Mevik B-H, Wehrens R, et al. pls: Partial Least Squares and Principal Component Regression. R package version 2.8–0. 2021.

30. Ministry of Health, Labour, and Welfare, Japan. Annual Report on Health, Labor and Welfare, 2016. https://www.mhlw.go.jp/wp/hakusyo/kousei/16/ (accessed 9 Mar 2022).

31. Organisation for Economic Co-operation and Development. Regions and Cities at a Glance 2018 – JAPAN. https://www.oecd.org/cfe/JAPAN-Regions-and-Cities-2018.pdf (accessed 9 Mar 2022).

32. Hara K, Kunisawa S, Sasaki N, et al. Future projection of the physician workforce and its geographical equity in Japan: a cohort-component model. BMJ Open 2018;8:e023696.

33. Matsuda S. Health Policy in Japan – Current Situation and Future Challenges. JMA J 2019;2:1–10.

34. Tanihara S, Kobayashi Y, Une H, et al. Urbanization and physician maldistribution: A longitudinal study in Japan. BMC Health Serv Res 2011;11.

35. Ramagem C, Urrutia S, Griffith T, et al. Combating health care fragmentation through integrated health services delivery networks. Int J Integr Care 2011;19.

36. Elrod JK, Fortenberry JL. The hub-and-spoke organization design: An avenue for serving patients well. BMC Health Serv Res 2017;17.

37. Alvarez Villela M, Clark R, William P, et al. Systems of Care in Cardiogenic Shock. Front Cardiovasc Med 2021;8:1–8.

